# Evaluating Redundancy and Biases in EHR Social Determinants of Health Data Screening

**DOI:** 10.64898/2026.02.18.26346575

**Authors:** John P. Powers, Amy Shaheen, Barbara Entwisle, Emily R. Pfaff

## Abstract

**Introduction:** Healthcare organizations have begun incorporating screening procedures for social determinants of health (SDOH) into care, recognizing the impact these factors can have on health outcomes. We aimed to present methods for evaluating redundancy in the risk information gained across SDOH questions and for evaluating whether demographic biases are present in whether patients were asked SDOH questions and whether they declined to answer them.

**Methods:** SDOH question data were analyzed for 1.8 million UNC Health patients. To evaluate risk information redundancy, response agreement was analyzed for pairs of questions. Demographic biases were evaluated using logistic regression models.

**Results:** Risk information redundancy was identified, particularly across food and financial insecurity questions. Furthermore, female and White patients were more likely to be asked some questions than other groups, and American Indian or Alaska Native and Hispanic or Latino patients were less likely to decline to answer questions.

**Conclusions:** We demonstrated methods healthcare organizations can use to evaluate their SDOH screening procedures. These methods yielded insights for (1) reducing burden in clinical workflows by identifying where redundancy could be eliminated and (2) reducing bias in SDOH data collection through more systematic screening protocols.

## INTRODUCTION

Social determinants of health (SDOH) have a significant impact on health, morbidity, and mortality.[1] With national health initiatives recognizing the import of SDOH in maintaining health,[2] governmental payers have begun requiring screening for SDOH in individual patients.[3] In anticipation of and in response to these requirements, many healthcare organizations have been rolling out SDOH screening procedures.[4–6] With substantial data now accumulated from these screening programs, organizations are seeking to evaluate the efficiency and effectiveness of SDOH screening programs to improve both their procedures and outcomes.[7,8] In this paper, we present analyses for evaluating efficiency and biases in SDOH screening at UNC Health, offering insights and replicable methods for evaluating and optimizing SDOH screening programs at other healthcare organizations.

The burden of adding several more questions to typical 15-minute visits or pre-visit questionnaires, as well as following up on the needs indicated by patients’ responses, can be a substantial concern for organizations trying to integrate SDOH screening practices.[4] This led us to query whether responses to some questions might be highly predictive of responses on other questions, i.e., the information captured by one question was redundant with another question. Such findings could support reducing the set of questions used in SDOH screening along with the burden of these screening procedures. For example, would financial resource strain accurately predict food insecurity or transportation barriers? While each SDOH question has been proven to predict its queried condition,[9–11] it was unknown whether responses in one domain could accurately predict responses in other domains. Thus, we used a data-driven approach to search for evidence of redundant risk information across SDOH questions, which could support eliminating questions to streamline screening procedures.

Additionally, the effectiveness of these screening procedures hinges on these questions being asked systematically across patients and patients choosing to answer these questions. Biases in either of these components could lead to patients with unmet social needs not being identified and appropriately supported. Indeed, SDOH screening rates have been shown to be associated with provider, patient, and setting characteristics as well as provider attitudes regarding SDOH screening, with effects sometimes varying by question domain.[12,13] Berg et al. have previously discussed potential sources of bias in SDOH data collected in electronic health records (EHR), including various selection biases and social biases influencing the data collected through direct interactions with providers.[14] Therefore, we evaluated SDOH response data for evidence of bias across demographic characteristics.

Thus, the first aim of these analyses was to identify redundancy in the risk information detected across the set of SDOH questions, which would support eliminating questions whose responses could be predicted from responses to the remaining questions. This would lower burden on practices and providers. Second, we aimed to evaluate whether biases were present between various demographic groups in (1) whether patients were asked SDOH questions during care and (2) whether patients declined to answer SDOH questions if asked. Such biases could indicate a need for developing more systematic SDOH screening procedures. We present these methods and findings to offer guidance to other institutions on how to evaluate and optimize SDOH data collection in their clinical care settings.

## MATERIALS AND METHODS

### Data

This study was approved by the University of North Carolina at Chapel Hill Institutional Review Board and received a waiver of informed consent for research and HIPAA authorization due to the research involving no greater than minimal risk. The data for this study were provisioned as part of a larger project examining SDOH in EHR at UNC Health, which started rolling out EHR SDOH questions and screening practices in late 2019. The dataset for the project included UNC Health EHR data in the Observational Medical Outcomes Partnership (OMOP) data model with a date range of Jan. 1, 2018– Dec. 31, 2022. In these initial years of SDOH screening rollout, the adoption of questions was highly variable across care sites, with care teams generally only being required to ask at least one SDOH question per patient per year. Thus, response rates were expected to be variable and generally low across all potential questions. Patients had to meet all of the following criteria for inclusion in the dataset: had a NC address in the date range, had at least two recorded encounters in the date range at least 30 days apart, had a recorded birth date, and was not incarcerated. The utilization inclusion criterion, two encounters at least 30 days apart, was used as a minimal indicator of active use of the healthcare system to filter out patients who may have only received care in the system for a single, isolated event. These analyses were additionally restricted to patients at least 18 years of age (N = 1,797,972). The dataset included SDOH question responses (see Table 1 for the list of questions and their labels) and demographic features. SDOH questions and answers were mapped to LOINC codes and stored in the OMOP data model using methods described in a prior publication.[15]

**Table 1.**
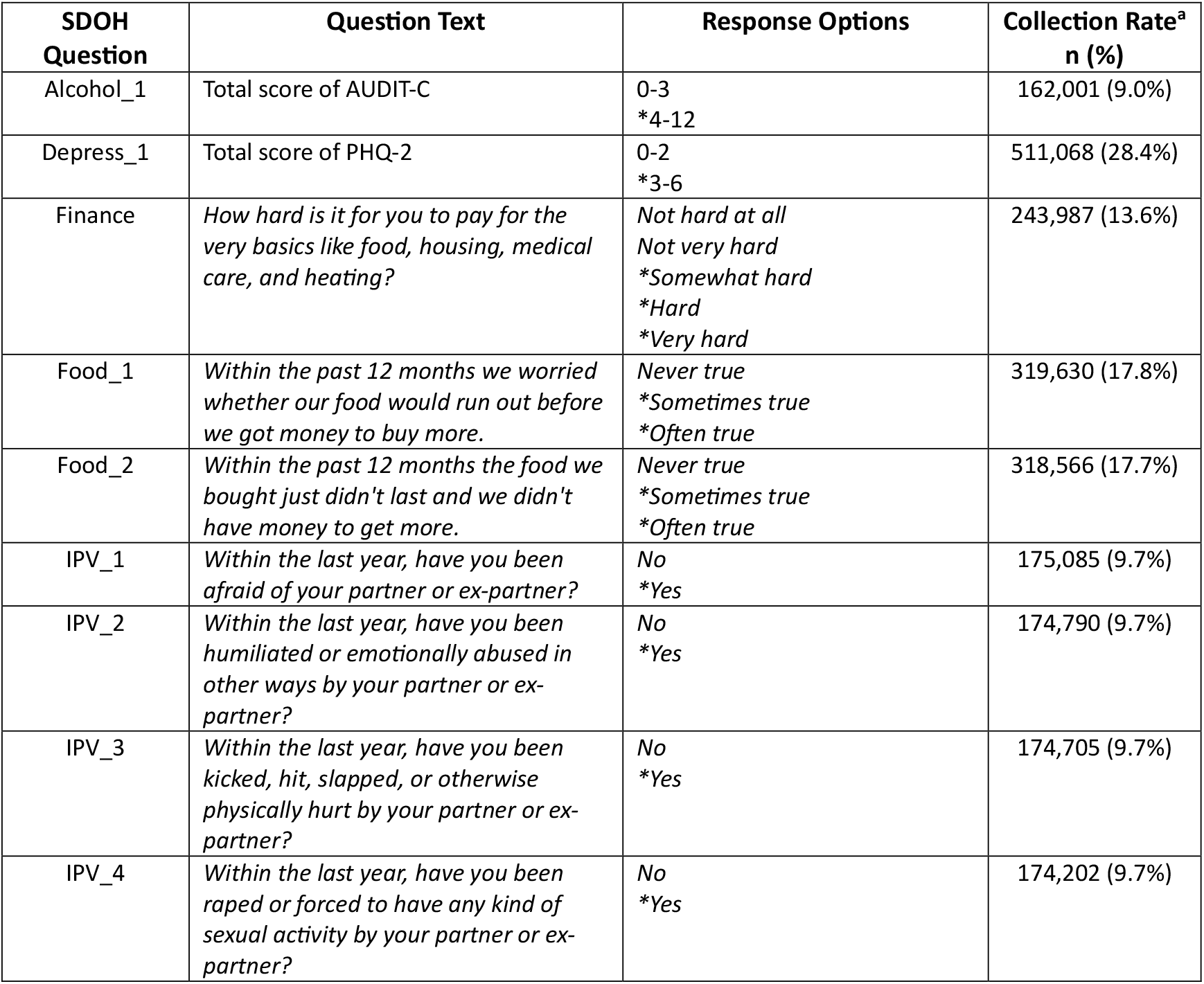
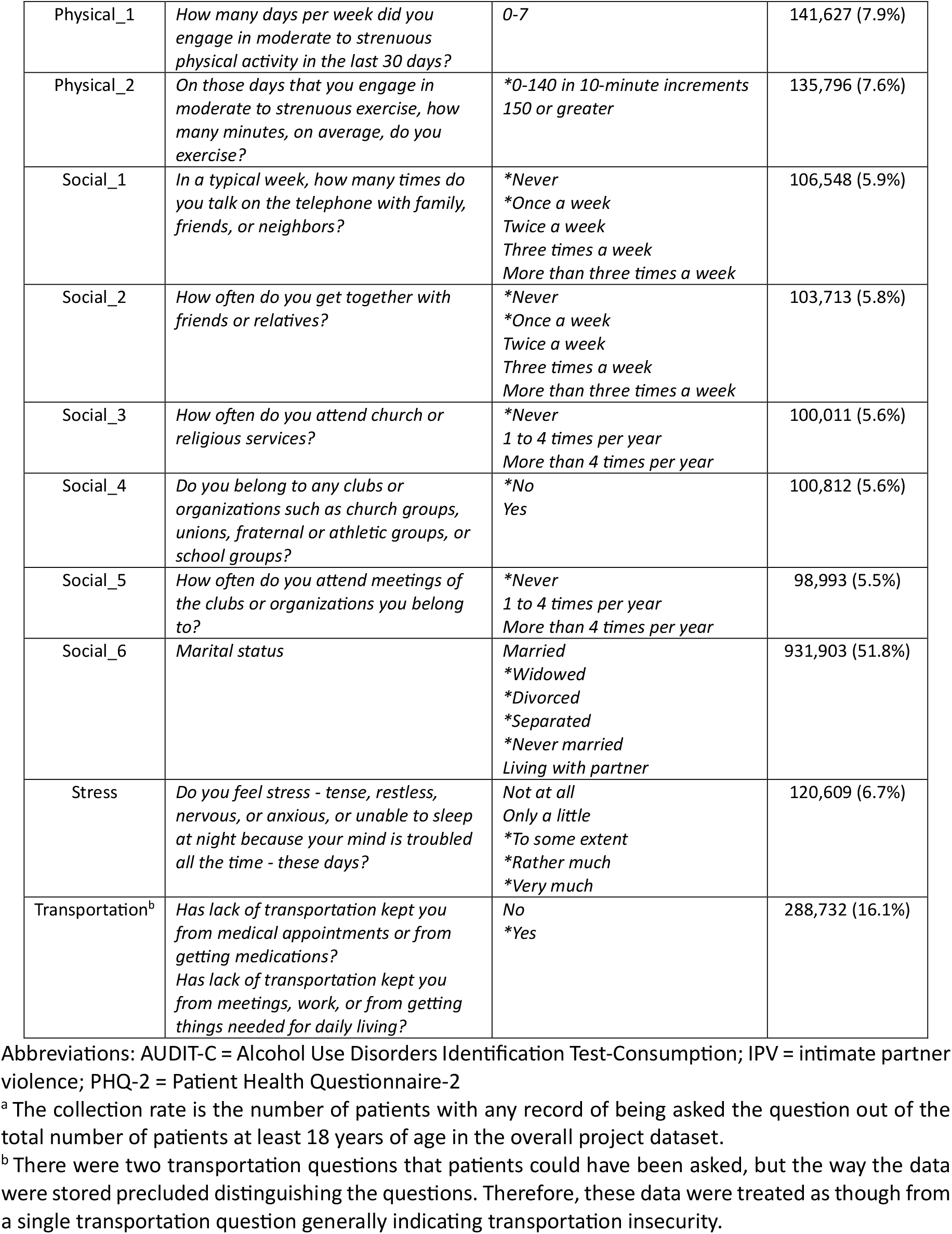
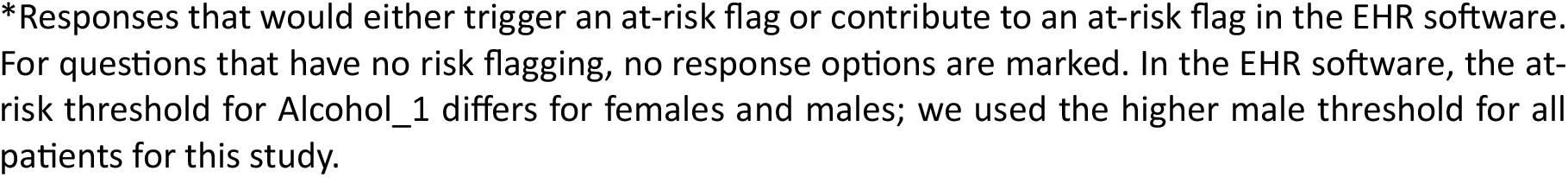
SDOH Questions.

| SDOH Question | Question Text | Response Options | Collection Rate<sup>a</sup><br>n (%) |
| --- | --- | --- | --- |
| Alcohol_1 | Total score of AUDIT-C | 0-3<br>*4-12 | 162,001 (9.0%) |
| Depress_1 | Total score of PHQ-2 | 0-2<br>*3-6 | 511,068 (28.4%) |
| Finance | *How hard is it for you to pay for the very basics like food, housing, medical care, and heating?* | *Not hard at all*<br>*Not very hard*<br>* *Somewhat hard*<br>* *Hard*<br>* *Very hard* | 243,987 (13.6%) |
| Food_1 | *Within the past 12 months we worried whether our food would run out before we got money to buy more.* | *Never true*<br>* *Sometimes true*<br>* *Often true* | 319,630 (17.8%) |
| Food_2 | *Within the past 12 months the food we bought just didn't last and we didn't have money to get more.* | *Never true*<br>* *Sometimes true*<br>* *Often true* | 318,566 (17.7%) |
| IPV_1 | *Within the last year, have you been afraid of your partner or ex-partner?* | *No*<br>* *Yes* | 175,085 (9.7%) |
| IPV_2 | *Within the last year, have you been humiliated or emotionally abused in other ways by your partner or ex-partner?* | *No*<br>* *Yes* | 174,790 (9.7%) |
| IPV_3 | *Within the last year, have you been kicked, hit, slapped, or otherwise physically hurt by your partner or ex-partner?* | *No*<br>* *Yes* | 174,705 (9.7%) |
| IPV_4 | *Within the last year, have you been raped or forced to have any kind of sexual activity by your partner or ex-partner?* | *No*<br>* *Yes* | 174,202 (9.7%) |
| Physical_1 | *How many days per week did you engage in moderate to strenuous physical activity in the last 30 days?* | *0-7* | 141,627 (7.9%) |
| Physical_2 | *On those days that you engage in moderate to strenuous exercise, how many minutes, on average, do you exercise?* | *\*0-140 in 10-minute increments*<br>*150 or greater* | 135,796 (7.6%) |
| Social_1 | *In a typical week, how many times do you talk on the telephone with family, friends, or neighbors?* | *\*Never*<br>*\*Once a week*<br>*Twice a week*<br>*Three times a week*<br>*More than three times a week* | 106,548 (5.9%) |
| Social_2 | *How often do you get together with friends or relatives?* | *\*Never*<br>*\*Once a week*<br>*Twice a week*<br>*Three times a week*<br>*More than three times a week* | 103,713 (5.8%) |
| Social_3 | *How often do you attend church or religious services?* | *\*Never*<br>*1 to 4 times per year*<br>*More than 4 times per year* | 100,011 (5.6%) |
| Social_4 | *Do you belong to any clubs or organizations such as church groups, unions, fraternal or athletic groups, or school groups?* | *\*No*<br>*Yes* | 100,812 (5.6%) |
| Social_5 | *How often do you attend meetings of the clubs or organizations you belong to?* | *\*Never*<br>*1 to 4 times per year*<br>*More than 4 times per year* | 98,993 (5.5%) |
| Social_6 | *Marital status* | *Married*<br>*\*Widowed*<br>*\*Divorced*<br>*\*Separated*<br>*\*Never married*<br>*Living with partner* | 931,903 (51.8%) |
| Stress | *Do you feel stress - tense, restless, nervous, or anxious, or unable to sleep at night because your mind is troubled all the time - these days?* | *Not at all*<br>*Only a little*<br>*\*To some extent*<br>*\*Rather much*<br>*\*Very much* | 120,609 (6.7%) |
| Transportation<sup>b</sup> | *Has lack of transportation kept you from medical appointments or from getting medications?*<br>*Has lack of transportation kept you from meetings, work, or from getting things needed for daily living?* | *No*<br>*\*Yes* | 288,732 (16.1%) |

Abbreviations: AUDIT-C = Alcohol Use Disorders Identification Test-Consumption; IPV = intimate partner violence; PHQ-2 = Patient Health Questionnaire-2
<sup>a</sup> The collection rate is the number of patients with any record of being asked the question out of the total number of patients at least 18 years of age in the overall project dataset.
<sup>b</sup> There were two transportation questions that patients could have been asked, but the way the data were stored precluded distinguishing the questions. Therefore, these data were treated as though from a single transportation question generally indicating transportation insecurity.
\*Responses that would either trigger an at-risk flag or contribute to an at-risk flag in the EHR software. For questions that have no risk flagging, no response options are marked. In the EHR software, the at-risk threshold for Alcohol\_1 differs for females and males; we used the higher male threshold for all patients for this study.

### Redundancy Analyses

#### Data Preprocessing

For many of the SDOH questions, thresholds have been programmed into the EHR software to flag a patient as being at-risk for negative health outcomes if their response is above the threshold (see Table 1). We used these thresholds to define a dichotomous risk status for these questions. For example, for Food_1 and Food_2, responses of *Never true* were categorized as not-at-risk, whereas responses of *Sometimes true* and *Often true* were categorized as at-risk. We eliminated duplicate records by filtering to the first record per question per patient per date. Through manual exploration, we observed such cases of multiple response records from a single date appeared to be duplicate data entries. If patients had multiple records for the same question collected on different dates, however, all of these records were retained in the dataset.

#### Analyses

Redundancy analyses involved evaluating agreement between responses recorded for the same patient and date for pairs of questions, with the goal of determining if the response to the first question in the pair was highly associated with the response to the second question. Thus, these analyses do not consider potential changes in patients’ responses to the same questions over time. Each pair of question responses from the same patient and date are considered independently, as the analysis is designed to assess the association between question responses that are collected concurrently. For all questions with a dichotomous risk status, the phi coefficient, which measures the strength of association between two binary variables, was computed between all possible question pairs. These phi coefficients were expressed as distance scores by subtracting them from one. A lower distance score then corresponds to a stronger association between measures. These distance scores were then submitted to hierarchical clustering analysis using the *linkage* function in the SciPy library for Python (v. 1.10.0) with the unweighted pair-group method with arithmetic mean. This hierarchical clustering analysis produces a summary of the relationships between all items based on all possible pairwise associations. Additional analyses were run on questions of interest (Finance, Food_1, Food_2, Transportation) in which all possible thresholds for dichotomizing the ordinal response options of one question in the pair were used to predict the original dichotomous risk status of the other question. Signal detection metrics were computed for each possible threshold of the predictor question, including sensitivity and specificity. A minimum sensitivity of 0.9 was used to identify significant predictions.

### Bias Analyses

#### Data Preprocessing

Data were limited to patients with recorded values for sex, race, and ethnicity. Although included in the redundancy analyses, data from patients identified as Native Hawaiian or Other Pacific Islander were excluded from declined/answered models (see below) due to insufficient counts for modeling.

#### Analyses

To evaluate biases in which patients were asked questions, we defined two groups for each question: (1) patients who were asked the question and (2) patients with no record of being asked the question but who received care since 2020 from the same sites where other patients were asked. This approach was used to help control for variance in SDOH screening by care site; only patients with records from care sites where SDOH questions were being asked were included in this analysis. Note, records could indicate that a patient declined to respond to a question; such a record was treated as an indication that the patient was asked the question. A logistic regression model was used to predict asked/not asked status for each question using age, sex, race, and ethnicity.

Similarly, to evaluate biases in which patients declined to answer a question when asked, we defined groups of patients who always declined the given question (this ranged from 0.6%-11.1% of patients, depending on the question) and patients who always answered the question. Patients with records of sometimes declining and sometimes answering the given question were excluded from this analysis (0.4%-1.8% of patients had this mixed pattern of responding, depending on the question). Note, a record of declining to answer a question was distinct from the absence of a record for that question; this analysis only involved records that explicitly indicated the patient declined to answer the question versus providing some other response. Logistic regression models predicted declined/answered status using age, sex, race, and ethnicity. The largest subgroup was used as the reference level for categorical predictors. Age was modeled as a continuous predictor. Only questions for which *declined* response data were present were included in these analyses.

Significant predictors met thresholds for statistical and practical significance. Statistical significance was defined as *p* < 0.05 for the Wald test. Practical significance was defined as an absolute change in log odds ratio greater than 0.223 (see “Practical Significance Threshold for Bias Analyses” in the Appendix for additional discussion of this threshold).

## RESULTS

Demographic characteristics of patients at least 18 years of age in the overall project dataset are summarized in Table 2.

**Table 2.** Demographic Characteristics of Patients.

| Characteristic | All Patients<sup>a</sup><br>n (%) | SDOH Patients<sup>b</sup><br>n (%) |
| --- | --- | --- |
| Total | 1,797,972 (100%) | 1,063,247 (100%) |
| Age |  |  |
| 18-19 | 40,576 (2.3%) | 26,274 (2.5%) |
| 20-29 | 230,190 (12.8%) | 144,612 (13.6%) |
| 30-39 | 252,185 (14.0%) | 155,631 (14.6%) |
| 40-49 | 255,405 (14.2%) | 149,941 (14.1%) |
| 50-59 | 301,095 (16.7%) | 173,081 (16.3%) |
| 60-69 | 314,963 (17.5%) | 185,051 (17.4%) |
| 70-79 | 249,718 (13.9%) | 145,525 (13.7%) |
| 80-89 | 117,470 (6.5%) | 65,604 (6.2%) |
| 90+ | 36,370 (2.0%) | 17,528 (1.6%) |
| Sex |  |  |
| Female | 1,068,161 (59.4%) | 620,172 (58.3%) |
| Male | 729,437 (40.6%) | 442,913 (41.7%) |
| Missing | 374 (<0.1%) | 162 (<0.1%) |
| Race |  |  |
| American Indian or Alaska Native | 33,440 (1.9%) | 23,729 (2.2%) |
| Asian | 36,189 (2.0%) | 24,267 (2.3%) |
| Black or African American | 362,965 (20.2%) | 229,274 (21.6%) |
| Native Hawaiian or Other Pacific Islander | 1,869 (0.1%) | 1,335 (0.1%) |
| White | 1,144,656 (63.7%) | 692,600 (65.1%) |
| Missing | 218,853 (12.2%) | 92,042 (8.7%) |
| Ethnicity |  |  |
| Hispanic or Latino | 116,866 (6.5%) | 68,868 (6.5%) |
| Not Hispanic or Latino | 1,576,334 (87.7%) | 967,538 (91.0%) |
| Missing | 104,772 (5.8%) | 26,841 (2.5%) |

<sup>a</sup> *All patients* refers to unique patients at least 18 years of age in the overall project sample (samples for each analysis varied based on specific analysis criteria and availability of the relevant data).
<sup>b</sup> *SDOH patients* refers to the subset of unique patients from *all patients* who were asked at least one of the social determinants of health questions listed in Table 1.

### Redundancy Analyses

Hierarchical clustering results are presented in Figure 1. We observed a very strong association between the two food questions (distance of 0.103), and a strong association between these food questions and the financial question (0.459). We also observed strong associations between intimate partner violence questions (distances between 0.306-0.594) and moderate-to-strong associations between social questions (0.567 for Social_1 and Social_2, 0.124-0.593 for Social_3-Social_5).

**Figure 1.**
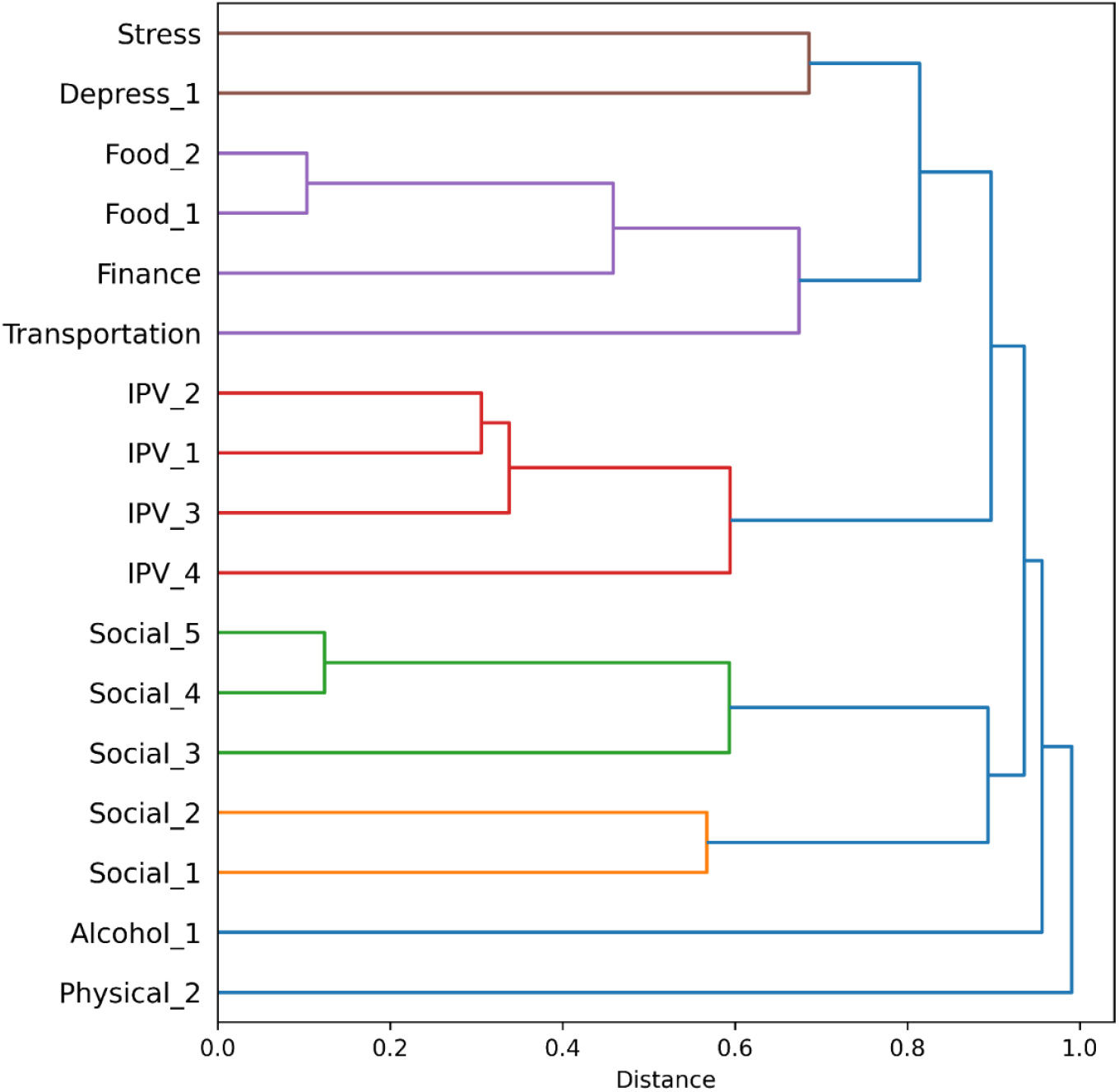
Clustering of questions based on associations between responses from patients. Clusters are colored based on a distance threshold of 0.7. Questions connected at shorter distances have more highly associated responses, such as Food_2 to Food_1 and Social_5 to Social_4.

Prediction analyses of questions of interest indicated that for one threshold of the financial question (*not hard at all* versus all other options), risk status for each food question was predicted with high sensitivity (Food_1: 0.912; Food_2: 0.921) but only modest specificity (Food_1: 0.760; Food_2: 0.719). However, risk status for Food_2 was predicted with high sensitivity (0.951) and specificity (0.993) by the original threshold of Food_1 (*never true* versus other options). No other predictions were significantly sensitive. This overall pattern of results replicated in several demographic subsamples (see “Redundancy Prediction Analyses in Subsamples” in the Appendix).

### Bias Analyses

Results for the bias analyses of asked/not asked and declined/answered (conditional on being asked) are summarized in Tables 3 and 4 respectively. For many questions, female and White patients were more likely to be asked than male patients and other non-Black or African American racial groups. Black or African American patients were more likely to be asked the financial and transportation questions when compared with White patients. American Indian or Alaska Native patients were less likely to decline to respond to questions than White patients, and Hispanic or Latino patients were less likely to decline to respond to questions than Not Hispanic or Latino patients. The likelihood of Asian patients declining to respond relative to White patients varied by question.

**Table 3.** Summary of Results for Asked/Not Asked Bias Analyses.

|  | Male<br>(vs. Female)<sup>a</sup> | AIAN | Asian<br>(vs. White) | Black | Pacific | Hispanic<br>(vs. Non-Hispanic) | Age |
| --- | --- | --- | --- | --- | --- | --- | --- |
| Finance | 0<sup>b</sup> | 0 | - | + | 0 | 0 | 0 |
| Food_1 | 0 | 0 | - | 0 | 0 | 0 | 0 |
| Food_2 | 0 | 0 | - | 0 | 0 | 0 | 0 |
| Transportation | 0 | 0 | 0 | + | 0 | 0 | 0 |
| Social_1 | - | - | - | 0 | - | 0 | 0 |
| Social_2 | - | - | - | 0 | - | 0 | 0 |
| Social_3 | - | - | - | 0 | - | 0 | 0 |
| Social_4 | - | - | - | 0 | - | 0 | 0 |
| Social_5 | - | - | - | 0 | - | 0 | 0 |
| Social_6 | - | - | - | 0 | - | 0 | 0 |
| IPV_1 | - | - | 0 | 0 | 0 | 0 | 0 |
| IPV_2 | - | - | 0 | 0 | 0 | 0 | 0 |
| IPV_3 | - | - | 0 | 0 | 0 | 0 | 0 |
| IPV_4 | - | - | 0 | 0 | 0 | 0 | 0 |
| Stress | - | - | - | 0 | - | 0 | 0 |
| Physical_1 | - | - | 0 | 0 | - | 0 | 0 |
| Physical_2 | - | - | 0 | 0 | - | 0 | 0 |

Abbreviations: AIAN = American Indian or Alaska Native; Black = Black or African American; Hispanic = Hispanic or Latino; IPV = intimate partner violence; Non-Hispanic = Not Hispanic or Latino; Pacific = Native Hawaiian or Other Pacific Islander
<sup>a</sup> Reference levels for categorical predictors are indicated in parentheses.
<sup>b</sup> Predictors with a significant negative effect on likelihood of being asked the question are indicated with a minus sign. Predictors with a significant positive effect on likelihood of being asked the question are indicated with a plus sign. Non-significant predictors are indicated with a zero.

**Table 4.** Summary of Results for Declined/Answered Bias Analyses.

|  | Male<br>(vs. Female)<sup>a</sup> | AIAN | Asian<br>(vs. White) | Black | Hispanic<br>(vs. Non-Hispanic) | Age |
| --- | --- | --- | --- | --- | --- | --- |
| Finance | 0<sup>b</sup> | - | + | 0 | - | - |
| Food_1 | 0 | - | 0 | 0 | - | 0 |
| Food_2 | 0 | - | 0 | 0 | 0 | 0 |
| Transportation | 0 | - | 0 | 0 | - | 0 |
| Social_1 | 0 | - | + | 0 | - | 0 |
| Social_2 | 0 | - | + | 0 | - | 0 |
| Social_3 | 0 | - | + | 0 | - | 0 |
| Social_4 | 0 | - | + | 0 | - | 0 |
| Social_5 | 0 | - | + | 0 | - | 0 |
| Social_6 | - | - | - | 0 | - | 0 |
| IPV_1 | 0 | - | - | 0 | - | 0 |
| IPV_2 | 0 | - | - | 0 | - | 0 |
| IPV_3 | 0 | - | - | 0 | - | 0 |
| IPV_4 | 0 | - | - | 0 | - | 0 |
| Stress | 0 | 0 | + | 0 | - | - |
| Physical_1 | 0 | 0 | 0 | 0 | - | - |
| Physical_2 | 0 | 0 | 0 | 0 | - | - |

Abbreviations: AIAN = American Indian or Alaska Native; Black = Black or African American; Hispanic = Hispanic or Latino; IPV = intimate partner violence; Non-Hispanic = Not Hispanic or Latino
<sup>a</sup> Reference levels for categorical predictors are indicated in parentheses.
<sup>b</sup> Predictors with a significant negative effect on likelihood of being asked the question are indicated with a minus sign. Predictors with a significant positive effect on likelihood of being asked the question are indicated with a plus sign. Non-significant predictors are indicated with a zero.

## DISCUSSION

This study aimed to evaluate whether multiple SDOH screening questions were providing redundant risk information and whether there were biases in which patients were asked SDOH questions and which patients declined to answer them. We demonstrated data-driven approaches using SDOH screening data that have been collected at UNC Health, and we found that the second of two food insecurity questions provided redundant information to the first and could therefore be dropped. This finding was replicated in several demographic subpopulations in addition to the full cohort. We also observed a few instances of moderately high redundancy (between food and finance questions, between multiple intimate partner violence questions, and between multiple social connection questions), where additional options for eliminating questions could be considered to further streamline screening procedures. At a minimum, these findings would support the removal of the highly redundant food question from the clinical workflow to reduce burden.

The definition of risk information used here involved applying thresholds to dichotomize ordinal responses into categories of at-risk or not-at-risk. While some response variance was sacrificed in this dichotomization, using thresholds to define actionable responses is often necessary to support clinical practice at scale, including in the context of SDOH screening.[16] For our initial redundancy analyses we relied on risk thresholds implemented in the EHR system (see Table 1 for how response options were dichotomized). For example, risk was considered positive for financial strain if a patient indicated “somewhat hard” or harder to pay for basics. Risk was considered positive for the food domain if a patient answered “sometimes true” or “often true” for questions about food running out. Some other SDOH domains, such as transportation and interpersonal safety, simply used dichotomous response options of yes or no. In an effort to support as many patients as possible, the healthcare system aimed for inclusive thresholds of positive risk to capture more patients who may benefit from intervention. Nevertheless, for select questions of interest we explored the full range of possible thresholds among response options.

We also found differences in rates of being asked and choosing to answer SDOH questions based on sex, race, and ethnicity. Whether one group is more or less likely to be asked or answer depends on the reference group. Here, where we used majority groups for reference, we found that male patients and patients of several non-White racial groups were less likely to be asked most questions than female and White patients. However, Black or African American patients were more likely to be asked the finance and transportation questions than White patients. We did not have data to determine the causes of these biases. Nevertheless, to minimize such biases in the future, we recommend that healthcare institutions provide clear, systematic guidelines for SDOH screening procedures, whether that means all patients are asked, or that a decision tree approach is used to determine who should be asked. The desired result is a policy that applies consistently to all patients.

One limitation of this dataset was the overall low collection rates of the SDOH questions. The rollout of SDOH screening at UNC Health has been a gradual process, as the adoption of new EHR tools often requires a significant transition period.[8] Current policies are increasing the number of SDOH questions patients are asked, but the study period of this dataset captured the initial rollout phase in which clinical teams were generally only required to ask patients at least one SDOH question. Thus, there may be biases not captured or addressed in these analyses in which questions care sites and providers selected to ask patients during the study period. The bias analyses presented here were designed to highlight broad patterns in SDOH screening across this health system. Steps were taken to reduce variance related to care site where relevant, as described in the methods. More granular factors, such as provider, likely impact SDOH data collection as well, but examination of these factors was beyond the scope of the present analyses. Additionally, the SDOH questions discussed in this study were designed to be collected as structured data in the EHR. Future research could explore supplementing such structured data with SDOH data extracted from clinical notes. This would require natural language processing tools to be feasible at this scale.[6]

In the context of brief clinical interactions, there is a tug-of-war between collecting maximal (and often regulatorily required) structured information and providing maximal care to the patient in the time allotted. New structured SDOH questions add to existing clinical processes, requiring more time to collect and follow up on. Based on these findings, we recommend careful evaluation of sets of screening questions to avoid collecting redundant risk information. We also recommend providing systematic guidelines for the clinic staff who ask SDOH questions and similar programming for automated screening systems to ensure screening procedures are implemented consistently across all patients. These recommendations and evaluation methods can be used to improve the efficiency and effectiveness of large-scale SDOH screening procedures at healthcare institutions.

## Data Availability

The analyses presented in this paper rely on a limited data set, as defined and protected by the Health Insurance Portability and Accountability Act Privacy Rule 45 CFR 164.514 issued by the U.S. Department of Health and Human Services and cannot be made publicly available.

## Appendix

### Practical Significance Threshold for Bias Analyses

Practical significance was defined as an absolute change in log odds ratio > 0.223 for one-hot encoded categorical variables, which included sex, race, and ethnicity. The same threshold for practical significance was used for age after multiplying its coefficient by 10 to reflect the change in log odds ratio for an age increase of 10 years rather than a single year. This threshold corresponds to an increase in odds ratio by a factor of 5:4 or a decrease in odds ratio by a factor of 4:5. For example, a negative effect in the Asian group on declining to respond would indicate the odds of declining to respond in the Asian group were less than the odds in the White group (the reference category) by a factor of at least 4:5. This threshold was selected to represent a minimal meaningful real-world difference in question asking or response patterns between demographic groups.

### Redundancy Prediction Analyses in Subsamples

The prediction analyses presented in the main text were also run in demographic subsamples for sex, race, and ethnicity. The significant results from the overall sample replicated in the female subsample, male subsample, Hispanic or Latino subsample, Not Hispanic or Latino subsample, White subsample, and Black or African American subsample. In the American Indian or Alaska Native subsample, Food_2 was not significantly predicted by Finance, but the other results replicated. Food_1 significantly predicted Food_2 in the Asian subsample, but the other results did not replicate. The subsample of Native Hawaiian or Other Pacific Islander was too small to meaningfully interpret. All replicated results noted above replicated using the same threshold of the predictor question as in the results from the main text.

We conclude that the findings of the redundancy prediction analyses in the main text are not driven by particular demographic subgroups; rather, they are consistent across most demographic groups. The cases in which the results did not replicate were in the smallest subsamples, where the effects may have been less stable due to smaller numbers. Even in these cases, the results that did not replicate were generally close to the significance threshold.

## References

1. CDC. Social determinants of health. Public Health Professionals Gateway. May 15, 2024. Accessed November 22, 2024. https://www.cdc.gov/public-health-gateway/php/about/social-determinants-of-health.html

2. Healthy People 2030. Accessed February 18, 2026. https://odphp.health.gov/healthypeople

3. Whitman A, De Lew N, Chappel A, Aysola V, Zuckerman R, Sommers BD. Addressing Social Determinants of Health: Examples of Successful Evidence-Based Strategies and Current Federal Efforts. Office of Health Policy, Office of the Assistant Secretary for Planning and Evaluation, U.S. Department of Health and Human Services; 2022. https://aspe.hhs.gov/sites/default/files/

4. Shaheen A, Squire MA, Gay H, Rana SY, Gwynne M, Gupta SK. System approaches to social determinants of health screening and intervention. NEJM Catal Innov Care Deliv. 2023;4(4). doi:10.1056/cat.22.0361

5. de la Vega PB, Losi S, Martinez LS, et al. Implementing an EHR-based screening and referral system to address social determinants of health in primary care. Med Care. 2019;57:S133–S139.

6. Li C, Mowery DL, Ma X, et al. Realizing the potential of social determinants data in EHR systems: A scoping review of approaches for screening, linkage, extraction, analysis, and interventions. J Clin Transl Sci. 2024;8(1):e147.

7. Wang M, Pantell MS, Gottlieb LM, Adler-Milstein J. Documentation and review of social determinants of health data in the EHR: measures and associated insights. J Am Med Inform Assoc. 2021;28(12):2608–2616.

8. Gold R, Bunce A, Cowburn S, et al. Adoption of social determinants of health EHR tools by community health centers. Ann Fam Med. 2018;16(5):399–407.

9. Hager ER, Quigg AM, Black MM, et al. Development and validity of a 2-item screen to identify families at risk for food insecurity. Pediatrics. 2010;126(1):e26–32.

10. Weir RC, Proser M, Jester M, Li V, Hood-Ronick CM, Gurewich D. Collecting social determinants of health data in the clinical setting: Findings from national PRAPARE implementation. J Health Care Poor Underserved. 2020;31(2):1018–1035.

11. Puterman E, Haritatos J, Adler NE, Sidney S, Schwartz JE, Epel ES. Indirect effect of financial strain on daily cortisol output through daily negative to positive affect index in the Coronary Artery Risk Development in Young Adults Study. Psychoneuroendocrinology. 2013;38(12):2883–2889.

12. Garg A, Cull W, Olson L, et al. Screening and referral for low-income families’ social determinants of health by US pediatricians. Acad Pediatr. 2019;19(8):875–883.

13. Wallace AS, Luther B, Guo JW, Wang CY, Sisler S, Wong B. Implementing a social determinants screening and referral infrastructure during routine emergency department visits, Utah, 2017-2018. Prev Chronic Dis. 2020;17(190339):E45.

14. Berg K, Doktorchik C, Quan H, Saini V. Automating data collection methods in electronic health record systems: a Social Determinant of Health (SDOH) viewpoint. Health Syst (Basingstoke). 2023;12(4):472–480.

15. Walters KM, Clark M, Dard S, et al. National COVID Cohort Collaborative data enhancements: A path for expanding common data models. J Am Med Inform Assoc. Published online November 23, 2024. doi:10.1093/jamia/ocae299

16. Billioux A, Verlander K, Anthony S, Alley D. Standardized screening for health related social needs in clinical settings: The Accountable Health Communities screening tool. NAM Perspect. Published online 2017. doi:10.31478/201705b

